# Rare and Common Genetic Variants, Smoking and Higher Body Mass Index Are Associated with Earlier Age of Progression to Geographic Atrophy and Neovascular Advanced Stages of Macular Degeneration in a Prospective Analysis

**DOI:** 10.1101/2020.08.13.20174383

**Authors:** Johanna M. Seddon, Rafael Widjajahakim, Bernard Rosner

**Affiliations:** Department of Ophthalmology and Visual Sciences, University of Massachusetts Medical School, Worcester, MA, USA; Channing Division of Network Medicine, Harvard Medical School, Boston, MA, USA

## Abstract

**IMPORTANCE:** Genes and lifestyle factors influence progression to advanced age-related macular degeneration (AAMD). However, the impact of genetic and behavioral factors on age when this transition occurs has not been evaluated prospectively.

**OBJECTIVE:** To determine whether genetic and environmental factors are associated with age of progression to AAMD and to quantify the effect on age.

**DESIGN, SETTING, AND PARTICIPANTS:** Longitudinal progression to AAMD was based on the severity scale in the Age-Related Eye Disease Study database. Progression was defined as an eye that transitioned from non-advanced dry AMD without any evidence of geographic atrophy (GA) (levels 1-8) to any GA or evidence of neovascularization (NV) or both (levels ≥9) during 13 years follow up. Genotypes were determined from DNA samples.

**MAIN OUTCOME AND MEASURES:** A stepwise selection of genetic variants with the eye as the unit of analysis, using age as the time scale, yielded 11 genetic variants associated with overall progression, adjusting for sex, education, smoking history, BMI, baseline severity scale, and AREDS treatment. Multivariate analysis was also performed to calculate the effect of genetic and behavioral factors on age of progression.

**RESULTS:** Among 5421 eyes, 1206 progressed. Genetic variants associated with progression to AAMD were in the complement, immune, inflammatory, lipid, extracellular matrix, DNA repair and protein binding pathways. Three of these variants were significantly associated with earlier age of progression, adjusting for other covariates: *CFH* R1210C (*P*=0.019) with 4.7 years earlier age at progression among carriers of this mutation, *C3* K155Q (*P*=0.011) with 2.44 years earlier for carriers, and *ARMS2/HTRA1* A69S (*P*=0.012) with 0.67 years earlier per allele. Subjects who were smokers (*P*<. 001) or had high BMI (*P*=0.006) also had an earlier age at progression (4.1 years and 1.4 years, respectively).

**CONCLUSIONS:** Carriers of rare variants in the complement pathway and a common risk allele in *ARMS2/HTRA1* develop advanced AMD at an earlier age, and unhealthy behaviors including smoking and higher body mass index lead to earlier age of progression to AAMD.

**KEY POINTS:** *Question:* Are genetic and modifiable factors associated with earlier age of developing advanced AMD?

*Findings:* In this prospective analysis of 5421 eyes with non-advanced AMD at baseline, 1206 developed advanced stages of AMD. Smoking together with a higher BMI led to 5.5 years earlier progression to advanced disease, and genetic burden, determined by rare and common variants, lowered age of progression by up to an additional 6.0 years, adjusting for all covariates.

*Meaning:* Modifiable factors alter age when advanced AMD and associated visual loss occurs, and genetic susceptibility impacts age of this transition. Results underscore the importance of both nature and nurture on earlier progression to advanced disease leading to a longer duration of disease and treatment burden. Submitted as ARVO abstract December, 2019: abstract citation: Widjajahakim R, Rosner B, Seddon J; Rare and Common Genetic Variants and Behavioral Modifiable Factors Are Associated with Earlier Age of Progression to Advanced AMD. *Invest. Ophthalmol. Vis. Sci*. 2020;61(7):2436).

## INTRODUCTION

Age-related macular degeneration (AMD) is a leading cause of vision loss and irreversible blindness in adults older than age 60, and has a complex etiology with both genes and environment contributing to risk.^1,2^ A set of genetic, demographic and environmental variables can predict with relatively high likelihood which individuals will be more likely to progress to AAMD.^3^ It has also been shown that individuals with rare genetic variants are more likely to progress^3,4^ and have an earlier diagnosis of AMD.^5^ However, the age at which the transition from non-advanced to AAMD occurs is variable, even among those with the same baseline macular pathology. The independent effect of individual variants and behavioral variables on this age of progression, and quantification of the potential number of years difference in age, have not been evaluated in a prospective study. We performed longitudinal analyses in a large, well defined cohort to assess the impact of both genetic and lifestyle factors on age when transition to advanced AMD occurs, adjusting for other known factors related to AMD. Results showed that rare and common genetic variants, smoking and higher body mass index were associated with earlier age of progression to geographic atrophy and neovascular stages of AMD in this prospective analysis.^6^

## METHODS

### Study Population and Classification of AMD Phenotypes

The Age-Related Eye Disease Study (AREDS) enrolled a total of 4,757 participants in the United States from 1992-1998.^7^ Among 6058 eyes, 5421 eyes with all covariate and genetic data were analyzed and 1206 eyes progressed to AAMD. The mean follow-up time was 9.3 years (range from 0.5 to 13 years). Progression to AAMD was based on the severity scale in the AREDS database.^8^ An eye that progressed was defined as transition from non-advanced dry AMD without any evidence of geographic atrophy (GA) (levels 1 -8) to any GA or evidence of neovascularization (NV) or both (levels ≥9) during 12 years follow up. The study adhered to the tenets of the Declaration of Helsinki and was performed under approved institutional review board protocols.

### Genotyping and Genetic Data

The DNA samples for the AREDS study population were purchased from the AREDS repository and genotyping was performed locally using array-based genotyping and gene sequencing platforms as previously described.^4,5,9-11^

### Statistical Methods

A stepwise selection was performed to determine which genetic variants among a set of previously reported variants related to AMD,^3^ were associated with progression from non-advanced to AAMD, adjusting for sex, education, smoking history, BMI, baseline severity scale, and AREDS treatment, using Cox proportional hazard regression models, with the eye as the unit of analysis and age as the time scale. The criteria for selection were *P*<0.05 for both entering and staying in the model. These stepwise regression models allow for the variables most predictive of a specific outcome to be determined based on an automatic procedure. Multivariate analysis was then performed to calculate the effect of each variant remaining in that model and behavioral covariates, on age of progression to AAMD using mixed effects regression models. For variables associated with earlier age of progression, histograms of mean age at progression were drawn. All analyses were calculated using SAS 9.4. Two-sided *P* values were calculated and 0.05 was used as the level of significance.

## RESULTS

A total of 11 out of 31 genetic variants were associated with progression to AAMD in the stepwise model (Table 1) adjusting for baseline severity scale and other covariates, representing several biologic pathways: complement, immune, inflammatory, lipid, extracellular matrix, DNA repair and protein binding. The estimated mean age of progression in the reference group (non-smoker, BMI<25, none of risk alleles, none of the risk factors) was 77.2 years (±2.2). Three of the 11 genetic variants showed a significant association with earlier age of progression to AAMD, adjusting for other covariates (Table 2): *CFH* R1210C: rs121913059 (*P*=0.019) with 4.7 years earlier age at progression among carriers of this mutation compared with non-carriers, *C3* K155Q: rs147859257 (P=.011) with 2.44 years earlier age at progression for carriers, and *ARMS2/HTRA1* A69S rs10490924 (*P*=.012) with 0.67 years earlier age of progression per allele. For carriers of the homozygous genotype for *ARMS2/HTRA1*, with 2 risk alleles, the impact would be 1.34 years earlier age of progression. Subjects who were current smokers (*P*<.001) or had higher BMI ≥30 (*P*=.006) also had an earlier age at progression to advanced disease (4.1years and 1.4 years, respectively), compared with the reference categories of never smoking or having a BMI < 25. The Figure displays some of these associations with the mean age of progression shifted toward younger ages for the rare *CFH* and *C3* rare variants, and the common *ARMS2/HTRA1* allele. Carrying the *CFH* R1210C mutation had the strongest genetic effect and current smoking had the largest behavioral impact on age of progression, adjusting for all covariates. Genetic and behavioral factors were independently related to earlier age of progression.

**Table 1.** Genetic Variants Associated With the Progression to Advanced AMD Using Stepwise Selection

| Genetic Variants | Estimates (±SE)<sup>a</sup> | P Value | HR (95% CI) |
| --- | --- | --- | --- |
| *CFH*: rs1410996 | 0.26 (±0.06) | <.001 | 1.30 (1.16-1.46) |
| *CFH* R1210C: rs121913059 | 0.99 (±0.26) | <.001 | 2.70 (1.64-4.48) |
| *C3* R102G: rs2230199 | 0.16 (±0.05) | <.001 | 1.18 (1.07-1.30) |
| *C3* K155Q: rs147859257 | 0.55 (±0.15) | <.001 | 1.74 (1.30-2.34) |
| *CFB* R32Q: rs641153 | -0.33 (±0.13) | .009 | 0.72 (0.56-0.92) |
| *ARMS2* A69S: rs10490924 | 0.32 (±0.05) | <.001 | 1.37 (1.26-1.51) |
| *COL8A1* P195L: rs13095226 | 0.16 (±0.07) | .023 | 1.17 (1.02-1.34) |
| *CETP*: rs3764261 | 0.10 (±0.05) | .032 | 1.11 (1.01-1.22) |
| *RAD51B*: rs8017304 | -0.18 (±0.05) | <.001 | 0.83 (0.75-0.92) |
| *TMEM97/VTN*: rs704 | -0.11 (±0.05) | .011 | 0.89 (0.82-0.97) |
| *PEL13* A307V: rs145732233 | -1.07 (±0.66) | .11<sup>b</sup> | 0.34 (0.09-1.26) |

<sup>a</sup> Estimates were calculated for time to progression using age as the time scale with eyes as the unit of analysis, adjusted for sex, education, smoking history, BMI, severity scale, and AREDS treatment. All genetic variants and non-genetic variables were analyzed in a single model. There were 1206 eyes which progressed among 5421 eyes. Requirement for both entering and staying in the model was $P < 0.05$ .
<sup>b</sup> *PEL13* was significant ( $P < 0.05$ ) based on a likelihood ratio test.

**Table 2.**
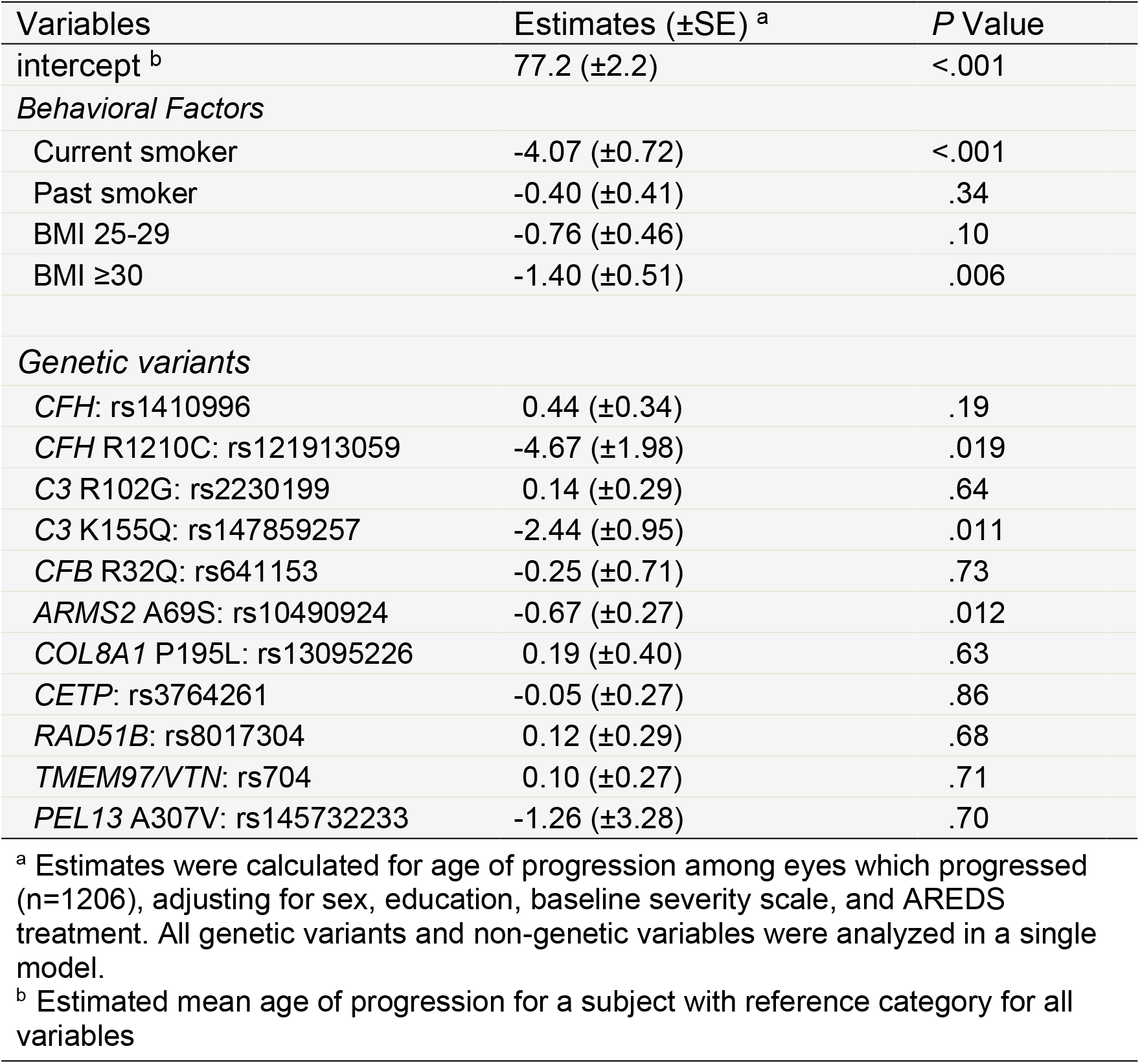
Multivariate Analysis of the Effects of Behavioral and Genetic Factors on Age of Progression to Advanced Age-Related Macular Degeneration

The combined effect of carrying the rare variant, *CFH* R1210C, plus two common *ARMS2/HTRA1* alleles, for an overweight patient who smoked, was progression to advanced AMD 11.5 years earlier, compared to not having this set of risk factors. Smoking and higher BMI alone without any of the genetic risk factors, conferred about a 5.5 year earlier onset of AAMD. Genetic burden, determined by the rare and common variants, lowered the age of progression by up to an additional 6.0 years.

## DISCUSSION

In this prospective analysis, we assessed the impact of genes and environment on age of progression to advanced AMD. Our results show that both nature and nurture influence when the late stages of AMD occur. Specifically, unhealthy behaviors, smoking and higher BMI,^12,13^ lead to earlier age of progression and thus a higher disease and treatment burden over several more years. Genetic susceptibility also plays a role, and higher genetic burden due to carrying high risk rare variants in *CFH* or *C3* in the complement pathway, or the common allele in the *ARMS2/HTRA1* gene, leads to earlier age of transitioning from non-advanced to advanced AMD. Of note, these genetic and behavioral factors were related to earlier disease burden independently, since analyses adjusted for all covariates including baseline demographic and ocular factors.

The literature regarding age of progression is sparse. A European study evaluated the neovascular form of AMD only in a cohort of 275 subjects in Europe, among which 214 had complete information, based on review of medical records in a retrospective study. They found that smoking history, *CFH* Y402H and the A69S variant in *ARMS2* were associated with earlier age of neovascular AMD.^14^ We have previously shown that carriers of *CFH* R1210C have younger age of onset of AMD in a case-control study^5^ and have higher risk of progression.^4^

The study reported herein differs in several ways: evaluation of a large cohort and prospective analysis of age of transition to both dry and exudative forms of advanced AMD, and inclusion of both rare and common genetic variants together with behavioral factors. We also adjusted the statistical model to account for correlation of eyes during the stepwise selection as well as all covariates in the same model to determine which variables were independently related. Other strengths include the standardized examinations and photographs at regular intervals, which more precisely determine age of progression. However, fundus photography was used to determine the severity scale and time of progression to advanced AMD, and optical coherence tomography could improve precision of these outcomes.^15^

### Conclusions

In summary, in this first prospective analysis of the impact of both genetic and non-genetic factors on age of progression from non-advanced to advanced AMD, results underscore the impact of both nature and nurture on developing advanced disease leading to visual loss at an earlier age. Smoking together with a higher BMI led to 5.5 years earlier progression to advanced disease, and genetic burden, determined by rare and common variants, lowered age of progression by up to an additional 6.0 years, leading to as much as 11.5 years earlier transition to advanced disease for genetic and behavioral risk factors combined, adjusting for all covariates. The serious impact of this earlier age of progression to advanced AMD with associated visual impairment is longer duration and burden of disease and treatment. Results of our analyses underscore the importance of adhering to healthy habits and provide biologic mechanisms to explore for preventive measures to shorten disease burden.

## Data Availability

Data is based on publicly available data from the U.S. National Institute of Health.

---

**FIGURE.**
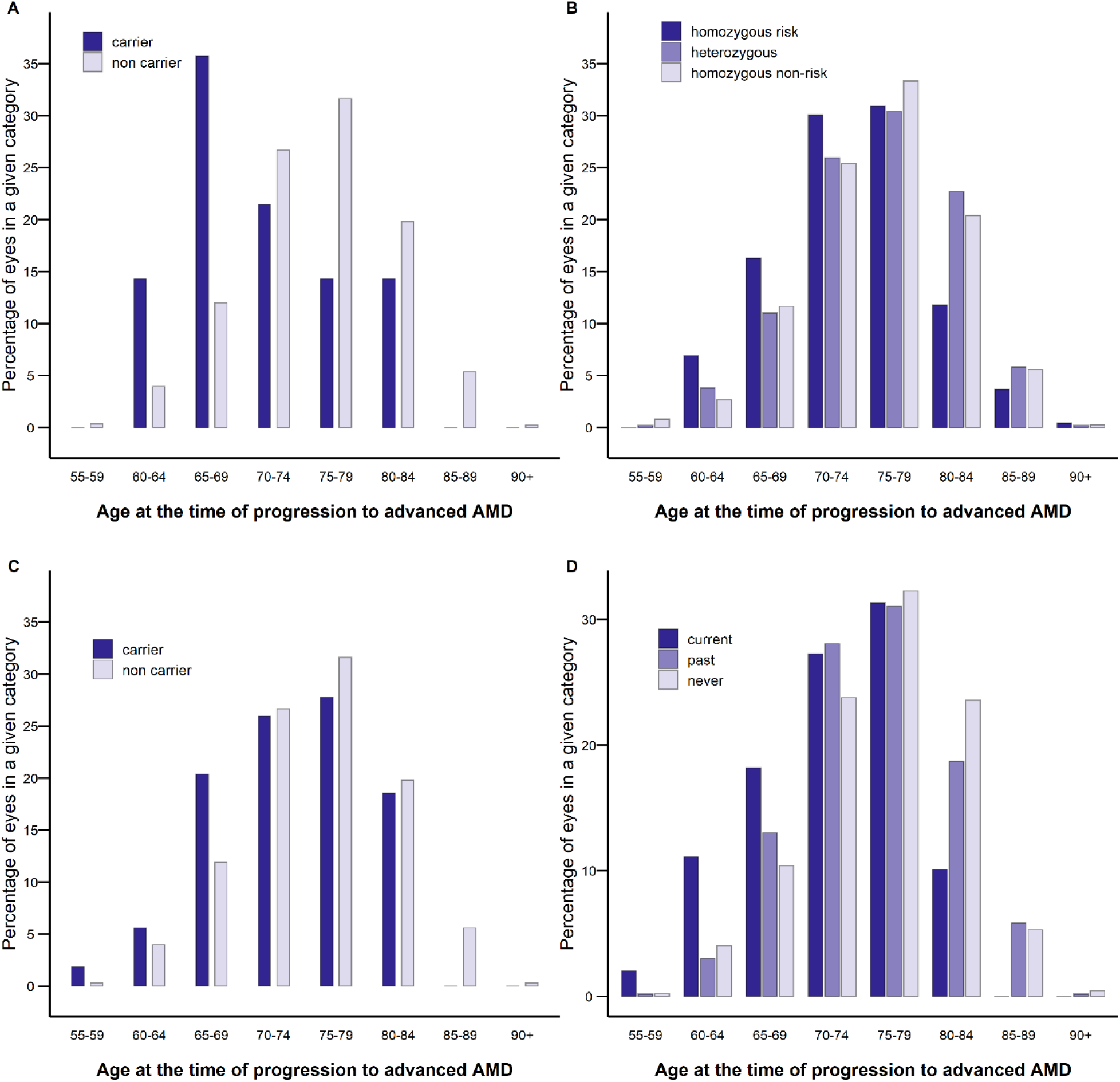
Age of progression to advanced AMD among progressors according to number of risk alleles for A) *CFH* R1210C, B) *ARMS2/HTRA1*, C) *C3* K155Q, and according to D) smoking status.

